# Working together effectively in research: Co-design and evaluation of capacity-building modules for researchers and people with lived experience of stroke

**DOI:** 10.1101/2025.07.07.25331049

**Authors:** Elizabeth A. Lynch, Stacy Larcombe, Julie Bernhardt, Brenda Booth, Liz Gallagher, Kathryn S Hayward, Gillian Mason, Adrian O’Malley, Ciara Shiggins, Dana Wong

**Affiliations:** College of Nursing and Health Sciences, Flinders University. Adelaide, Australia; NHMRC Centre for Research Excellence in Stroke Rehabilitation and Brain Recovery, Melbourne, Australia; Florey Institute of Neuroscience and Mental Health, University of Melbourne, Melbourne, Australia; Independent Lived Experience Expert, Woy Woy, Australia; Stroke Foundation, Melbourne, Australia; Departments of Physiotherapy and Medicine, University of Melbourne, Melbourne, Australia; Consumer and Community Involvement Team, Hunter Medical Research Institute, Newcastle, Australia; Independent Lived Experience Expert, Sydney, Australia; NHMRC Centre of Research Excellence in Aphasia Recovery and Rehabilitation, Melbourne, Australia; School of Health and Rehabilitation Sciences, The University of Queensland, Brisbane, Australia; Surgical Treatment and Rehabilitation Service (STARs), The University of Queensland and Metro North Health, Brisbane, Australia; School of Psychology and Public Health, La Trobe University, Melbourne, Australia

**Keywords:** Stroke, Patient and Public Involvement, Co-Design, Education, Evaluation

## Abstract

**Objectives:** To co-design, co-produce and evaluate resources to build the capacity of stroke researchers and people with lived experience of stroke to work together effectively on research projects.

**Methods:** Following an interactive workshop, a team of health professional researchers, people with lived experience of stroke, and an education specialist from Stroke Foundation (Australia) convened to co-design and co-produce two learning modules. Mixed methods were used to evaluate the modules. Researchers were invited to complete a survey about their capacity, motivation and intended behaviour before and after accessing the module. Researchers and people with lived experience who accessed the modules were invited to participate in interviews. Survey data were analysed descriptively and pre-and post-module responses were compared using t-tests. Interview data were analysed using qualitative content analysis by three researchers and one survivor of stroke.

**Results:** The modules have been widely accessed (researcher module n=1,115 users from 16 countries, lived experience module n=119 users on 20^th^ May 2025). Forty-one researchers completed surveys. Twelve researchers (n=9, 75% women; n=9, 75% from Australia) and 11 people with lived experience of stroke (n=6, 55% women; n=11, 100% from Australia) participated in interviews. Most participants had previous experience of collaborative research. Despite this, participants from both cohorts described a better understanding of roles and types of involvement after completing the modules. Researchers reported significant increases in their likelihood of involving lived experience contributors in research processes after completing the module, compared to responses pre-module.

**Conclusion:** Two learning modules co-designed and co-produced by health professional researchers, people with lived experience of stroke, and Stroke Foundation representatives led to improvements in knowledge and confidence for stroke researchers and those with lived experience of stroke to engage in collaborative research. Modules are freely available and compulsory for all researchers submitting Stroke Foundation grant applications.

## 1. Introduction

There has been a consistent push internationally to improve the way healthcare users are involved in, and inform, health research.(1) Terminology used to describe this involvement varies between regions and includes patient and public involvement,(2) public and patient engagement,(3) consumer and community engagement,(4) knowledge-user or patient engagement,(5, 6) co-creation, co-design and co-production.(7) National research funding bodies in many countries including Australia, USA, Canada and the UK require applicants to outline how people living with the clinical condition under investigation, or people who support them (e.g. families, informal carers), hereafter referred to collectively as “people with lived experience”, are involved in the proposed research.(3, 4, 8, 9) However, many people with research qualifications (hereafter called “researchers”) and people with lived experience do not feel confident to work collaboratively in research.(10, 11)

Researching in partnership, collaboration or consultation with people with lived experience requires researchers with clinical backgrounds to shift from ‘health professional-to-patient’ mindsets that are traditionally associated with a power differential,(12) towards peer or collegial relationships(13) that recognise and acknowledge the value of lived experience and resultant expertise. The COM-B model indicates that an individual’s behaviour (B in the model) is reliant on their skills and confidence (Capability, C), resources and social factors such as culture (Opportunity, O) and their motivation (M).(14) Applying this model, researchers need the skills and confidence, appropriate resources, a supportive research culture, and the motivation (influenced by both personal values and external incentives) to conduct research “with” people with lived experience. Similarly, for people with lived experience to be involved in research in ways other than as research participants (hereafter referred to as “lived experience contributors”), they too need skills, confidence, the desire/motivation to be involved, adequate time and energy, and necessary adaptations and accommodations to be able to contribute effectively. Aligning with the theory that building capacity supports behaviour change, there has been a steady increase in generic publications and resources about how best to work with people with lived experience and support their involvement in research.(15–17)

Condition-specific resources are likely required to meet the needs and context of discrete populations. For example, common post-stroke sequalae include communication difficulty (affecting 54% of stroke survivors),(18) cognitive changes (affecting up to 60% of survivors),(19) depression and anxiety (affecting 30% of survivors),(20) fatigue (affecting 48% of survivors)(21) and physical disabilities such as difficulty walking (affecting 80% of survivors).(22)

This paper outlines work conducted in Australia to co-design, co-produce and evaluate resources to build the capacity of people with lived experience of stroke and researchers to work together effectively in research.

The research aims were to:

- Co-design and co-produce educational resources to enhance the capacity of researchers to work effectively with people with lived experience of stroke.
- Co-design and co-produce educational resources to enhance the capacity of people with lived experience of stroke to work effectively with researchers.
- Evaluate whether module completion led to a change in capability, opportunity, motivation and/or intended behaviour in researchers and people with lived experience of stroke to work together on research projects.

## 2. Methods

### 2.1 Development of learning modules

To explore ways to promote diversity and inclusion in stroke research, the NHMRC Centre of Research Excellence in Stroke Rehabilitation and Brain Recovery (CRE-Stroke) convened a workshop in Melbourne, Australia, in February 2020. Health professionals, academic researchers and people with lived experience of stroke (survivors and people in their support network) were invited to attend. Travel assistance was provided to attendees with lived experience. The workshop was co-facilitated by a health professional researcher (author EAL) and a survivor of stroke with extensive advocacy experience (author AO’M).

Workshop activities comprised presentations, facilitated discussions and small group activities to strategize how to support researchers and people with lived experience of stroke to work together effectively in research. One priority identified was the need for training resources to support researchers and people with lived experience of stroke to work together. Funding was subsequently provided by Stroke Foundation (Australia) and CRE-Stroke to create learning modules.

Following the workshop, recordings and notes were reviewed by the authorship team, which comprised health professional researchers (n=6), people with lived experience of stroke (n=2) and an education coordinator from Stroke Foundation (n=1). Workshop materials, evidence from the literature and authors’ experiences of involvement in stroke research were integrated to create and refine educational content for the modules, with monthly online meetings held between October 2020 and July 2021. The team developed two draft modules (one for researchers working in the field of stroke, another for people with lived experience of stroke) that were shared with attendees of the 2020 workshop to review and provide feedback, which was then incorporated into the modules. Authors CS (speech and language therapist) and LG (education coordinator) formatted the module for people with lived experience using aphasia friendly principles(23), adding features such as a read-aloud function to enhance accessibility.

The module Working effectively with people with lived experience to design, conduct and promote stroke research(24) was released via the Stroke Foundation InformMe website (designed for health professionals and researchers) in March 2022. The module comprised 23 interactive slides with six videos which presented the perspectives of researchers and lived experience contributors, different ways to involve people with lived experience.

Learning opportunities were supported by knowledge checks and links to resources.

The module Working well with stroke researchers(25) was released via the Stroke Foundation EnableMe website (designed for people with lived experience of stroke) in April 2022. This module consisted of 36 slides with five videos which presented the perspectives of researchers and lived experience contributors about the value of involvement and potential roles in research. Both modules provided an overview of the research cycle, a downloadable “Top tips for collaboration” checklist, and links to additional resources.

Both modules were advertised in the Stroke Foundation newsletter and social media channels (Facebook, Instagram, and X [formerly Twitter]). Presentations and workshops about the resources were delivered at conferences and special interest groups in Australia, UK, and New Zealand, and to online international audiences between 2021 and 2024. The “Top tips” for collaborating with survivors of stroke were re-formatted for publication as a tutorial for Early Career Researchers and trainees.(26)

### 2.2 Evaluating the resources

Mixed methods, comprising surveys and interviews, were used to evaluate the learning modules. Ethical approval was granted by the Flinders University Human Research Ethics Committee (project number:4718).

Researchers who logged into the Working effectively with people with lived experience…module were invited to complete a survey before and after accessing the training module. On completion of the module, users were invited to provide their contact details if they were willing to be interviewed about the module.

People with lived experience of stroke, aged 18 and over, who completed the Working well with stroke researchers module were invited to provide their contact details if they were willing to be interviewed about the module.

#### 2.2.1 Data collection

##### 2.2.1.1 Survey

The authorship team developed a survey for researchers who accessed the training module (see supplementary file). The decision not to develop a survey for people with lived experience was advised by the lived experience team-members. Responses to questions about researchers’ intended behaviour were rated on a 5-point Likert scale (not-at-all-likely to very-likely). Confidence and satisfaction questions were rated on a 10-point Likert scale (not-at-all-confident/satisfied to extremely-confident/satisfied). Survey data were collected between February 2022 and October 2023.

##### 2.2.1.2 Interviews

Interview guides comprising open-ended questions were developed by the authorship team for the two participant groups (See supplementary file). Question development was guided by the COM-B model for Behaviour Change(14) to understand whether the module influenced how respondents conducted or were involved in research through changing capability, opportunity or motivation. The interview guides were pilot tested for appropriateness and to ensure attainment of quality data. No changes were required.

Individuals who expressed interest in being part of the interviews were emailed information sheets and consent forms. When possible, group interviews (up to three participants) were scheduled with researchers, but if scheduling was problematic, individual interviews were organised. All participants with lived experience of stroke were interviewed individually.

All interviews were conducted via videoconferencing (Microsoft Teams, video-recorded) or via phone (audio-recorded) between March and October 2023 by authors EAL and SL. Interviews were transcribed verbatim and de-identified for confidentiality purposes.

#### 2.2.2 Data analysis

Survey data were analysed descriptively. For survey questions asked before-and-after module completion, repeated measures t-tests were used to analyse changes in responses, with effect sizes calculated using Cohen’s d.

Interview data were analysed using qualitative content analysis(27). Two authors (SL, EAL) read the transcripts and identified codes pertinent to the research questions (whether completing the module influenced the respondent’s knowledge, skills, confidence, motivation or intentions to involve/be involved in research). Data from researchers were analysed separately from data from lived experience contributors. Interview summaries were provided to participants to review for accuracy.

Codes were transferred to IdeaFlip(28) for the second stage of analysis by authors EAL, SL, BB. Each code was presented on a virtual sticky note, and exemplary quotes were presented on the reverse of each note. Author BB (a lived experience contributor) reviewed the coded data but had not read the transcripts. Codes were defined and refined through online discussions, and transcripts were reviewed when the context of a quote required clarification. Each code was given a short description, and categorised as Capability (e.g. skills, knowledge), Opportunity (e.g. resources, culture), Motivation or Behaviour (working with lived experience contributors). Author DW read through the organised codes and provided feedback on the way they had been categorised within the framework.

Analyses from surveys and interviews were tabled. When available, data from interviews were used to illustrate or expand on survey findings.

## 3. RESULTS

### 3.1 Website use

As of 20^th^ May 2025, the researcher module was accessed by 1,115 users from 16 countries, and the lived experience module by 119 users from Australia.

### 3.2 Participant characteristics

Forty-one researchers completed surveys. Most respondents were women (n=33, 80%), university-based (n=29, 71%), aged between 25-45 years (n=24, 59%) with a health professional background (n=31, 76%).

Twelve researchers (n=9, 75% women) participated in interviews. Nine researchers were based in Australia, with one researcher each from Canada, Italy and the USA. Experience working in the field of stroke (as a clinician or as a researcher) ranged from less than one year to 20 years. Eleven people with lived experience of stroke (n=6, 55% women) participated in interviews. All were in Australia with a time since stroke between 1 and 10 years. Most interview participants (researchers n=10, 83%; lived experience n=8, 72%) had experience working on research projects that involved people with lived experience.

### 3.3 Module evaluations

Results from the survey and interviews are presented according to the main categories of the COM-B model. Survey responses are presented in Table 1. Quotes from interview participants are annotated as researcher (R) or person with lived experience of stroke (PWLE).

**Table 1:**
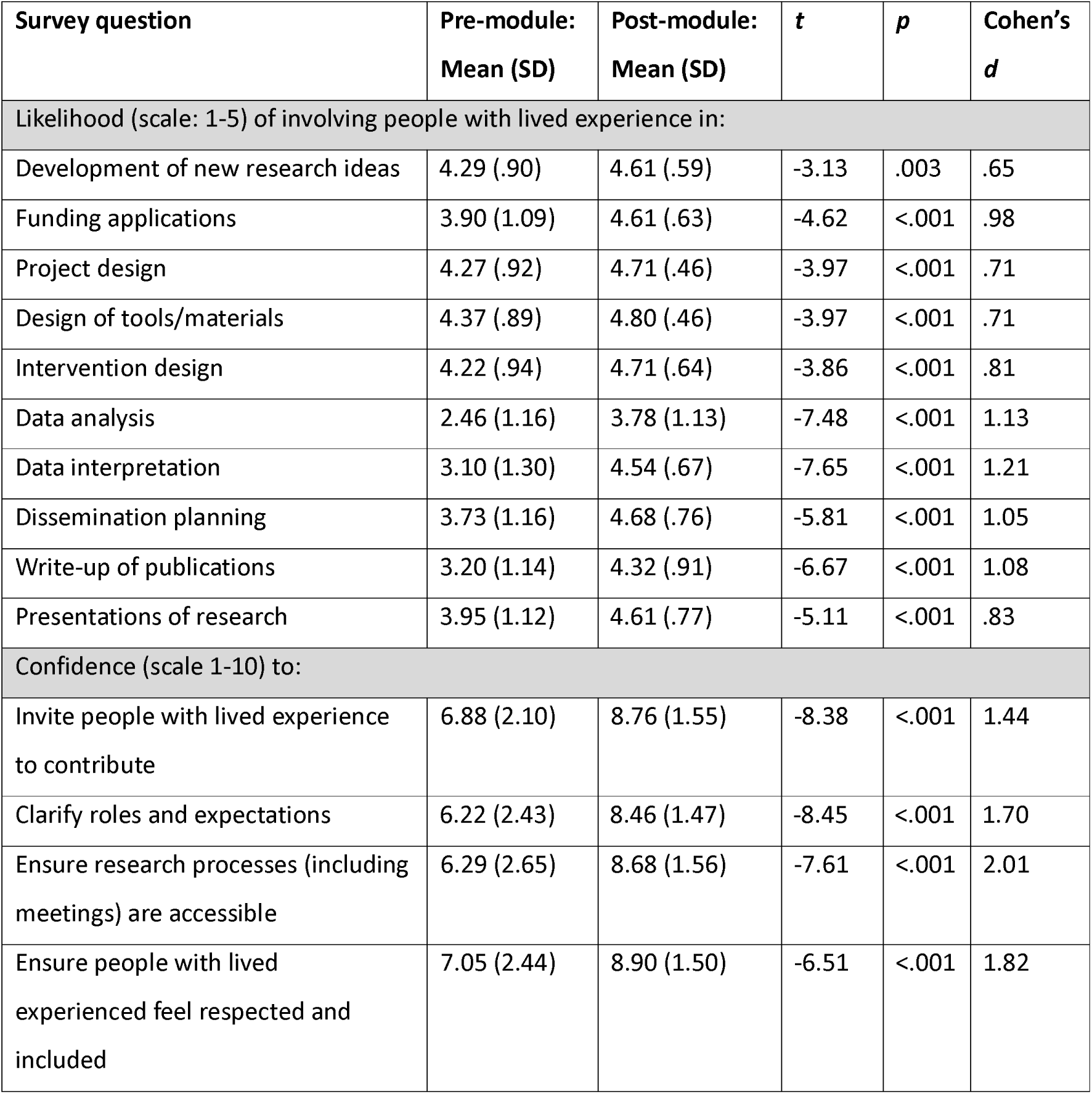
Descriptive statistics and comparisons between pre-and post-module survey questions (n=41)

| Survey question | Pre-module:<br>Mean (SD) | Post-module:<br>Mean (SD) | *t* | *p* | Cohen's<br>*d* |
| --- | --- | --- | --- | --- | --- |
| Likelihood (scale: 1-5) of involving people with lived experience in: |  |  |  |  |  |
| Development of new research ideas | 4.29 (.90) | 4.61 (.59) | -3.13 | .003 | .65 |
| Funding applications | 3.90 (1.09) | 4.61 (.63) | -4.62 | <.001 | .98 |
| Project design | 4.27 (.92) | 4.71 (.46) | -3.97 | <.001 | .71 |
| Design of tools/materials | 4.37 (.89) | 4.80 (.46) | -3.97 | <.001 | .71 |
| Intervention design | 4.22 (.94) | 4.71 (.64) | -3.86 | <.001 | .81 |
| Data analysis | 2.46 (1.16) | 3.78 (1.13) | -7.48 | <.001 | 1.13 |
| Data interpretation | 3.10 (1.30) | 4.54 (.67) | -7.65 | <.001 | 1.21 |
| Dissemination planning | 3.73 (1.16) | 4.68 (.76) | -5.81 | <.001 | 1.05 |
| Write-up of publications | 3.20 (1.14) | 4.32 (.91) | -6.67 | <.001 | 1.08 |
| Presentations of research | 3.95 (1.12) | 4.61 (.77) | -5.11 | <.001 | .83 |
| Confidence (scale 1-10) to: |  |  |  |  |  |
| Invite people with lived experience to contribute | 6.88 (2.10) | 8.76 (1.55) | -8.38 | <.001 | 1.44 |
| Clarify roles and expectations | 6.22 (2.43) | 8.46 (1.47) | -8.45 | <.001 | 1.70 |
| Ensure research processes (including meetings) are accessible | 6.29 (2.65) | 8.68 (1.56) | -7.61 | <.001 | 2.01 |
| Ensure people with lived experienced feel respected and included | 7.05 (2.44) | 8.90 (1.50) | -6.51 | <.001 | 1.82 |

#### 3.3.1 Capability

##### 3.3.1.1 Strengthening Knowledge and Skills (interview data)

People with lived experience reported the module “Working well with stroke researchers” was delivered at an introductory level and was useful for building knowledge and competence. Participants from both groups consistently reported that completion of the modules led to a better understanding of the types and levels of involvement in research, and how roles could be defined for the different levels of involvement.

> “The research cycle is useful. I probably wouldn’t have known about the research cycle and the levels of involvement or engagement of people with lived experience”[R3]

> “So now I’ve got a picture of the whole life cycle of the research project”[PWLE6]

The modules assisted participants to clarify the rationale for involvement and presented a “clearer path”[PWLE5] for collaboration. Researchers and lived experience contributors acknowledged the modules enhanced insight into “the value of lived experience”[PWLE8] and highlighted that a person with lived experience is “an expert in the experience of stroke”.[R10] Further, respondents reported that the modules highlighted the skills that people with lived experience of stroke could bring to a project from both their stroke and other life experiences.

Researchers reported that their module was helpful for learning how to describe the different levels of involvement. Preferred terminology to describe the people with whom they worked was also deemed useful.

> “The module gave me…extra language to use in terms of like a lived experience contributor or partner.”[R10]

> “Whether it is a partnership or a consultation”[R3].

Content and practical strategies to proactively seek guidance on how to adjust ways of working to meet a lived experience contributor’s needs were beneficial for some researcher participants.

> “Someone might be getting to the end of their tolerance for sitting up, engaging, conversing and cognitively switching on for that conversation to really keep to time so I’ve kept that in mind as a practical thing”[R10]

Multi-media presentation of information, such as the infographic of the research cycle, and real-life examples presented in video format, were frequently reported to enhance participants’ understanding of how researchers and people with lived experience could work together. Adjoining links and regular knowledge checks throughout the researcher module were reported as effective learning strategies. Respondents reported that the downloadable checklist summarised the key module components and was a useful resource for future projects.

Lived experience contributors suggested development of a subsequent Lived Experience module presenting more technical detail on research design, team composition and strategies to support involvement in research. Suggested additions to the module for researchers included incorporating examples of administrative documents such as Terms of Reference and position descriptions for lived experience contributors, stroke-specific documents to meet the needs of people with cognitive or language difficulties, and guidance to navigate reimbursement and reporting of involvement of lived-experience contributors.

##### 3.3.1.2 Building Confidence (survey and interview data)

Survey responses indicated researchers were more confident in their skills after completing the module, particularly regarding clarifying roles and expectations, making research processes accessible, and ensuring lived experience contributors feel respected and included (see Table 1). These responses echoed interview findings. Participants with lived experience described more confidence to work collaboratively. Participants from both groups felt more confident to ask questions to ensure both parties were working effectively together.

> “More confidence to know where or what my contribution might be…not to be so reticent…for giving my opinions on things.”[PWLE6]

> “I feel more confident…in knowing that there will be bits that are hard and…just being confident to ask the person with lived experience how to do it right and how to work with them correctly.”[R8]

One researcher reported that the module boosted their confidence in their research skills, because the module reinforced that their ways of working were consistent with recommended practice. Endorsement of the module by Stroke Foundation appeared to increase the trustworthiness of the content, thereby adding to the researcher’s confidence.

> “[The module] definitely made me feel more confident to [involve people with lived experience]…if I hadn’t done the module, I think I still would have done it…effectively and successfully. But I felt more confident having…this Stroke Foundation tick of approval…I certainly felt more confident because it’s the Stroke Foundation’s module”.[R7]

#### 3.3.2 Opportunity (survey and interview data)

The value of the module as a resource for researchers was reflected in high satisfaction scores in the survey (mean rating 8.6 for the content of the module, 8.5 for its presentation, 9 for its usefulness in informing research practice). All survey respondents (100%) said they would recommend the module to other researchers in the stroke field. This feedback was also reflected in interviews, with both groups reporting that the modules were valuable resources that were easy to use, with clearly presented content. The time taken to complete the module (generally less than 30 minutes) was deemed appropriate by both participant groups. Researchers appreciated the ability to return and review module content, although the log-in requirement to access the researcher module was identified as a potential barrier to its use. Several researchers reported planning to use the module to guide future projects.

> “I’ll definitely look at the module again before doing a research project”[R8]

Elements within the modules, particularly the Top Tips and case examples were deemed helpful to enhance learning and to support future research projects.

> “The thing that like the ten steps that you could think about…that is really important”[PWLE10]

> “When the training module got into the actual examples…it was really great to see the examples of what…stroke research might look like and how lived participants can contribute to the design of it”[PWLE4]

> “I liked the steps for how to engage and define roles from the outset”[R8]

One researcher described the module as a vehicle for changing research culture, by guiding and upskilling more researchers and people with lived experience to enable future research collaborations.

> “Some of the benefits of having toolkits like this is that…all of us who are trying to continue this culture of including and understanding the value of people’s experience and we can use this toolkit to…speak to the masses…so that people entering this space understand that this is this is how you do it.…there’s no other way to do research now in stroke, this is the way”[R3]

A suggested change to the researcher module to improve opportunity was to embed a system to facilitate connections with lived experience contributors.

#### 3.3.3 Motivation (interview data)

Generally, participants reported accessing and completing the modules because they were already motivated to work collaboratively with researchers or people with lived experience of stroke and were looking to advance their skills and knowledge. In these instances, motivation to work collaboratively was already strong and did not change following completion of the module. Because of this, participants were also asked about their motivations to complete the module. Motivation for researchers to complete the module and to work collaboratively with people with lived experience both appeared to be influenced by research funding bodies. Many researchers mentioned the requirement from funding bodies to demonstrate how people with lived experience are involved in the research, and several researchers reported completing the module due to the Stroke Foundation Research Partnership Framework mandate, wherein completion of the co-designed module was required for applicants to be considered for Stroke Foundation grant funding.(29)

> “It’s become increasingly important in grants, writing and anything, any kind of funding that you’re applying for that…you have to show where you’ve at least thought about…consumer involvement…I didn’t know what that meant and was looking for more information about that”[R4]

> “My honest answer is that I was going to apply for a Stroke Foundation grant and it said you need to do the module”[R8]

#### 3.3.4 Behaviour (survey and interview data)

Researchers who completed the surveys indicated they were more likely to involve lived experience contributors in research processes after completing the module, compared to pre-module (all t-tests comparing pre-post responses were statistically significant, p<0.01, see table 1). Effect sizes were medium-large, with particularly large increases on questions regarding intentions to involve lived experience contributors in data analysis, interpretation, and dissemination.

In interviews, one researcher reported changing research practices as a direct result of completing the module, by inviting people with lived experience of stroke to collaborate on a grant application and modifying ongoing research protocols to enhance the involvement of lived-experience contributors in research projects. Several researchers indicated that they intended to change their behaviours and were more likely to partner with lived-experience contributors on future projects. Further, researchers reported considering different types and levels of involvement and planned to use the checklist provided in the module to ensure true involvement and engagement of people with lived experience so that “the efforts that are made to include stroke survivors are not tokenistic”[R10].

## 4. Discussion

### 4.1 Discussion

We have described how we co-designed, co-produced and evaluated two learning modules to enhance involvement in stroke research. The modules were conceptualised at a workshop coordinated and attended by researchers, people with lived experience of stroke and health professionals. Deliberate efforts were made to optimise accessibility of the module for stroke survivors, including those with cognitive and/or communication challenges. The module for researchers also included specific content on how to involve people with communication support needs throughout the research process. Evaluation methods were guided by recommendations from team members with lived experience. Data were analysed by three researchers and one lived experience contributor. The resources have been disseminated by researchers and lived experience contributors. By working in this collaborative way from conceptualisation through to dissemination of findings, our team has sought to do as we say, “work effectively together in research”.

We found that completion of the learning modules improved self-reported skills and confidence of researchers and people with lived experience of stroke to involve or be involved in research. This is an important national initiative because Australia currently lags behind the UK and Canada in terms of consistency of involving people with lived experience in research.(30) Other Australian initiatives specific to stroke, beyond work described in this manuscript, include the development of resources to support better inclusion of people with communication difficulties as both participants and as contributors in research (CRE-Aphasia).(31) These purpose-designed and freely available resources can action behaviour change both through improving access to tools and educational resources, as well as through the social influence of the people who lead these initiatives, when these people are respected and trusted.(32)

Beyond changing an individual’s behaviour through training and education, organizational systems and policies can also affect change.(14) Mandates from Stroke Foundation (Australia) for completion of the learning module by all researchers seeking to partner with, or apply for funding from them(29) will ensure that more Australian researchers access information about involving people with lived experience in research. Mandates from national funders to describe how people with lived experience will be involved in the research will similarly encourage all researchers seeking funding to strive towards involvement. However, further policy-level changes could be of benefit to address structural barriers that influence research practices. A common barrier, that could be overcome with organisational support is the scarcity of funding available to reimburse lived experience contributors for their time, especially when they are supporting grant-writing, when funding is not yet available and may not be awarded.(33)

As research involvement becomes more commonplace, the research community needs to create systems that recognise these changes. Human research ethics committees have long been stalwarts of promoting safe, just and respectful research,(34) but application and review processes are not always conducive to involving people with lived experience in research.(35) Our team has heard frequent anecdotal reports, which are also reported in the published literature,(35) of ethics review committees requesting that people with lived experience who are involved as consultants or advisors in research projects sign participant consent forms, thereby being inappropriately considered as participants. Further, ethics approvals can be difficult when methods are yet to be co-designed, such as in participatory action research.(36) Changing ethical review processes is complex, but important for advancing involvement in research as well as healthcare delivery.(37)

The strength of this work is the robust collaboration between researchers and those with lived experience throughout. Limitations to our evaluation include that all participants were already involved or interested in collaborative research practices and we cannot know whether the modules would be of value in changing the perceptions of individuals who are less interested in working collaboratively.

### 4.2 Conclusion

We have described a successful project wherein a team of researchers and people with lived experience of stroke have co-designed and co-produced two educational modules to support capacity building in researchers and people with lived experience of stroke regarding involvement practices. The modules have been accessed widely. Users reported that accessing the modules improved their skills and knowledge to work collaboratively on research projects.

### 4.3 Practice implications

To advance involvement of people with lived experience in research, researchers and people with lived experience need appropriate knowledge, skills and confidence, supportive environments and social networks and the desire to work together. Addressing knowledge and skills through developing training resources is one important strategy. For sustainability, initiatives addressing individuals’ knowledge and skills should be accompanied by strategies addressing systems, such as practical, structural and appropriately resourced support provided by the academic or healthcare institution.

## Data Availability

All data produced in the present study are available upon reasonable request to the authors

## Acknowledgements

Sincere thanks and gratitude to workshop attendees for their time, valuable insights, experiences, and feedback throughout the co-design process and to individuals who made videos for the training modules. The authors would also like to thank the NHMRC Centre of Research Excellence in Stroke Rehabilitation Recovery and Stroke Foundation for financial support to develop the modules. KSH is funded by NHMRC Investigator Award (2016420) and Heart Foundation Future Leader Fellowship (106607).

## REFERENCES

1. Aiyegbusi OL, McMullan C, Hughes SE, Turner GM, Subramanian A, Hotham R, et al. Considerations for patient and public involvement and engagement in health research. Nat Med. 2023;29:1922–9.

2. National Institute for Health and Care Research (NIHR). PPI (Patient and Public Involvement) resources for applicants to NIHR research programmes: Department of Health and Social Care; 2019 [Available from: https://www.nihr.ac.uk/documents/ppi-patient-and-public-involvement-resources-for-applicants-to-nihr-research-programmes/23437. Accessed 30^th^ May 2025

3. Patient-Centered Outcomes Research Institute (PCORI). Public and Patient Engagement Washington, DC: Patient-Centered Outcomes Research Institute (PCORI); 2018 [Available from: https://www.pcori.org/about/about-pcori/our-programs/engagement/public-and-patient-engagement. Accessed 30^th^ May 2025

4. National Health and Medical Research Council (NHMRC). Consumer and community engagement 2021 [Available from: https://www.nhmrc.gov.au/about-us/consumer-and-community-involvement/consumer-and-community-engagement. Accessed 30^th^ May 2025

5. Canadian Institutes of Health Research (CIHR). Knowledge User Engagement: Government of Canada; 2016 [Available from: https://cihr-irsc.gc.ca/e/49505.html. Accessed 30^th^ May 2025

6. Canadian Institutes of Health Research (CIHR). Patient engagement: Government of Canada; 2019 [Available from: https://cihr-irsc.gc.ca/e/45851.html. Accessed 30^th^ May 2025

7. Vargas C, Whelan J, Brimblecombe J, Allender S. Co-creation, co-design, co-production for public health – a perspective on definitions and distinctions. Public Health Res Pract. 2022;15:32:3222211

8. National Institute for Health and Care Research. Briefing notes for researchers - public involvement in NHS, health and social care research: NIHR; 2021 [Available from: https://www.nihr.ac.uk/documents/briefing-notes-for-researchers-public-involvement-in-nhs-health-and-social-care-research/27371. Accessed 30^th^ May 2025

9. Canadian Institutes of Health Research. Strategy for Patient-Oriented Research: Government of Canada; 2023 [Available from: https://cihr-irsc.gc.ca/e/41204.html. Accessed 30^th^ May 2025

10. Boylan AM, Locock L, Thomson R, Staniszewska S. “About sixty per cent I want to do it”: Health researchers’ attitudes to, and experiences of, patient and public involvement (PPI)-A qualitative interview study. Health Expect. 2019;22:721–30.

11. Agyei-Manu E, Atkins N, Lee B, Rostron J, Dozier M, Smith M, et al. The benefits, challenges, and best practice for patient and public involvement in evidence synthesis: A systematic review and thematic synthesis. Health Expect. 2023;26:1436–52.

12. Nimmon L, Stenfors-Hayes T. The “Handling” of power in the physician-patient encounter: perceptions from experienced physicians. BMC Med Educ. 2016;16:114.

13. Farr M. Power dynamics and collaborative mechanisms in co-production and co-design processes. Crit Soc Policy. 2018;38:623–44.

14. Michie S, van Stralen MM, West R. The behaviour change wheel. A new method for characterising and designing behaviour change interventions Implementation Sci. 2011;6:42.

15. Harrison JD, Auerbach AD, Anderson W, Fagan M, Carnie M, Hanson C, et al. Patient stakeholder engagement in research: A narrative review to describe foundational principles and best practice activities. Health Expect. 2019;22:307–16.

16. Smits DW, van Meeteren K, Klem M, Alsem M, Ketelaar M. Designing a tool to support patient and public involvement in research projects: the Involvement Matrix. Res Involv Engagem. 2020;6:30.

17. Ocloo J, Garfield S, Franklin BD, Dawson S. Exploring the theory, barriers and enablers for patient and public involvement across health, social care and patient safety: a systematic review of reviews. Health Res Policy Syst. 2021;19:8.

18. De Cock E, Batens K, Hemelsoet D, Boon P, Oostra K, De Herdt V. Dysphagia, dysarthria and aphasia following a first acute ischaemic stroke: incidence and associated factors. Eur J Neurol. 2020;27:2014–21.

19. El Husseini N, Katzan IL, Rost NS, Blake ML, Byun E, Pendlebury ST, et al. Cognitive Impairment After Ischemic and Hemorrhagic Stroke: A Scientific Statement From the American Heart Association/American Stroke Association. Stroke. 2023;54:e272–91.

20. Chun H-YY, Ford A, Kutlubaev MA, Almeida OP, Mead GE. Depression, Anxiety, and Suicide After Stroke: A Narrative Review of the Best Available Evidence. Stroke. 2022;53:1402–10.

21. Alghamdi I, Ariti C, Williams A, Wood E, Hewitt J. Prevalence of fatigue after stroke: A systematic review and meta-analysis. Eur Stroke J. 2021;6:319–32.

22. Moore SA, Boyne P, Fulk G, Verheyden G, Fini NA. Walk the Talk: Current Evidence for Walking Recovery After Stroke, Future Pathways and a Mission for Research and Clinical Practice. Stroke. 2022;53:3494–505.

23. Rose TA, Worrall LE, Hickson LM, Hoffmann TC. Aphasia friendly written health information: content and design characteristics. Int J Speech Lang Pathol. 2011;13:335–47.

24. Stroke Foundation. Working effectively with people with lived experience to design, conduct and promote stroke research 2022 [Available from: https://informme.org.au/learning-modules/working-effectively-with-people-with-lived-experience-to-design-conduct-and-promote-stroke-research. Accessed 30^th^ May 2025

25. Stroke Foundation. Working well with stroke researchers 2022 [Available from: https://enableme.s3.ap-southeast-2.amazonaws.com/Research/story.html. Accessed 30^th^ May 2025

26. Lynch EA, Booth B, O’Malley A, Hayward KS, Mason G, Shiggins C, et al. How to Work Effectively With Stroke Survivors Throughout the Research Process. Stroke. 2024;55:e258–e61

27. Hsieh H-F, Shannon SE. Three approaches to qualitative content analysis. Qual Health Res. 2005;15:1277.

28. BIGGERFLIP LTD. Online Sticky Notes 2024 [Available from: https://ideaflip.com/. Accessed 30^th^ May 2025

29. Stroke Foundation. Stroke Foundation Research Grants 2024 [Available from: https://strokefoundation.org.au/what-we-do/research/research-grants.

30. Lang I, King A, Jenkins G, Boddy K, Khan Z, Liabo K. How common is patient and public involvement (PPI)? Cross-sectional analysis of frequency of PPI reporting in health research papers and associations with methods, funding sources and other factors. BMJ Open. 2022;12:e063356.

31. La Trobe University. Centre for Research Excellence in Aphasia Recovery and Rehabilitation Melbourne, Australia [Available from: https://www.latrobe.edu.au/research/centres/health/aphasia. Accessed 30^th^ May 2025

32. Rogers EM, Singhal A, Quinlan MM. Diffusion of innovations. An integrated approach to communication theory and research: Routledge; 2014. p. 432–48.

33. Foster A, Caunt S, Schofield H, Glerum–Brooks K, Begum S, Gleeson P, et al. Evaluating a grant development public involvement funding scheme: a qualitative document analysis. Res Involv Engagem. 2024;10:57.

34. Rhodes R. Rethinking Research Ethics. Am J Bioeth. 2005;5:7–28.

35. Nollett C, Eberl M, Fitzgibbon J, Joseph-Williams N, Hatch S. Public involvement and engagement in scientific research and higher education: the only way is ethics? Res Involv Engagem. 2024;10:50.

36. Goodyear-Smith F, Jackson C, Greenhalgh T. Co-design and implementation research: challenges and solutions for ethics committees. BMC Med Ethics. 2015;16:78.

37. Glasziou P, Scott AM, Chalmers I, Kolstoe SE, Davies HT. Improving research ethics review and governance can improve human health. J R Soc Med. 2021;114:556–62.

