## Supplementary table for "Working together effectively in research: Co-design and evaluation of capacity-building modules for researchers and people with lived experience of stroke"

**Supplementary File 1. Questions for quantitative survey**

**PRE-TRAINING:**

Q1 What is your current work or study setting? (select all that apply)

- University
- Hospital/inpatient setting
- Outpatient/community setting
- Other:

Q2 What is your age?

- 18-24
- 25-34
- 35-44
- 45-54
- 55-64
- 65+

Q3 What is your gender?

- Female
- Male
- Other
- Prefer not to say

Q4 What is your professional background/qualification/area of study? (select all that apply):

- Clinical/Health Psychology
- Clinical Neuropsychology
- Medicine. Please add specialty: ____________________________________
- Nurse
- Nutrition/Dietetics
- Occupational Therapy
- Paramedicine/Ambulance service
- Pharmacy
- Physiotherapy
- Social work
- Speech Pathology
- Other: (10) ________________________________________________

Q5: How many years of research experience do you have? (Please include years completing PhD/Doctorate): ________________

Q6: Please mark the research activities where you think it is important to work with people with lived experience:

- Identifying/developing a new research idea
- Applying for funding
- Designing a project
- Designing a data collection tool (e.g. a test, questionnaire or interview questions)
- Designing an intervention
- Analysing or interpreting data
- Planning the dissemination strategies
- Writing publications
- Presenting research results

Q7: Please mark the research activities where you have previously worked with people with lived experience:

- Identifying/developing a new research idea
- Applying for funding
- Designing a project
- Designing a data collection tool (e.g. a test, questionnaire or interview questions)
- Designing an intervention
- Analysing or interpreting data
- Planning the dissemination strategies
- Writing publications
- Presenting research results

Please rate your agreement with the following statements

| Q8 | I understand what is involved when working with people with lived experience on a research projects | | | | |
| --- | --- | --- | --- | --- | --- |
|  | - Strongly disagree | - Strongly disagree | - Neither agree nor disagree | - Agree | - Strongly agree |
| Q9 | I am confident that I can clarify roles and expectations with lived experience contributors | | | | |
|  | - Strongly disagree | - Strongly disagree | - Neither agree nor disagree | - Agree | - Strongly agree |
| Q10 | I am able to ensure research processes, including meetings, are accessible to people with mobility difficulties | | | | |
|  | - Strongly disagree | - Strongly disagree | - Neither agree nor disagree | - Agree | - Strongly agree |
| Q11 | I am able to ensure research processes and meetings are accessible to people with cognitive or fatigue difficulties | | | | |
|  | - Strongly disagree | - Strongly disagree | - Neither agree nor disagree | - Agree | - Strongly agree |
| Q12 | I am able to ensure research processes and meetings are accessible to people with communication difficulties | | | | |
|  | - Strongly disagree | - Strongly disagree | - Neither agree nor disagree | - Agree | - Strongly agree |
| Q13 | I know other researchers who have worked with people with lived experience on research projects | | | | |
|  | - Strongly disagree | - Strongly disagree | - Neither agree nor disagree | - Agree | - Strongly agree |
| Q14 | There are systems in place to support me to work with people with lived experience | | | | |
|  | - Strongly disagree | - Strongly disagree | - Neither agree nor disagree | - Agree | - Strongly agree |
| Q15 | Working with people with lived experience is not part of my role as a researcher | | | | |
|  | - Strongly disagree | - Strongly disagree | - Neither agree nor disagree | - Agree | - Strongly agree |
| Q16 | Working with people with lived experience would be easy | | | | |
|  | - Strongly disagree | - Strongly disagree | - Neither agree nor disagree | - Agree | - Strongly agree |
| Q17 | Working with people with lived experience would not be of benefit | | | | |
|  | - Strongly disagree | - Strongly disagree | - Neither agree nor disagree | - Agree | - Strongly agree |
| Q18 | There are no incentives to work with people with lived experience on research projects | | | | |
|  | - Strongly disagree | - Strongly disagree | - Neither agree nor disagree | - Agree | - Strongly agree |
| Q19 | Working on a research project with people with lived experience would make me feel uncomfortable | | | | |
|  | - Strongly disagree | - Strongly disagree | - Neither agree nor disagree | - Agree | - Strongly agree |
| Q20 | I have adequate resources (e.g. time, money) to work with people with lived experience on my research projects | | | | |
|  | - Strongly disagree | - Strongly disagree | - Neither agree nor disagree | - Agree | - Strongly agree |
| Q21 | Working with people with lived experience on my research projects would be viewed well by my peers | | | | |
|  | - Strongly disagree | - Strongly disagree | - Neither agree nor disagree | - Agree | - Strongly agree |

**POST-TRAINING**

Q1: Please mark the research activities you think you would be likely to work with people with lived experience within the next 12-18 months:

- Identifying/developing a new research idea
- Applying for funding
- Designing a project
- Designing a data collection tool (e.g. a test, questionnaire or interview questions)
- Designing of an intervention
- Analysing or interpreting data
- Planning the dissemination strategies
- Writing publications
- Presenting research results

**Please rate your agreement with the following statements**

| Q2 | I understand what is involved when working with people with lived experience on a research projects | | | | |
| --- | --- | --- | --- | --- | --- |
|  | - Strongly disagree | - Strongly disagree | - Neither agree nor disagree | - Agree | - Strongly agree |
| Q3 | I am confident that I can clarify roles and expectations with lived experience contributors | | | | |
|  | - Strongly disagree | - Strongly disagree | - Neither agree nor disagree | - Agree | - Strongly agree |
| Q4 | I am able to ensure research processes, including meetings, are accessible to people with mobility difficulties | | | | |
|  | - Strongly disagree | - Strongly disagree | - Neither agree nor disagree | - Agree | - Strongly agree |
| Q5 | I am able to ensure research processes and meetings are accessible to people with cognitive or fatigue difficulties | | | | |
|  | - Strongly disagree | - Strongly disagree | - Neither agree nor disagree | - Agree | - Strongly agree |
| Q6 | I am able to ensure research processes and meetings are accessible to people with communication difficulties | | | | |
|  | - Strongly disagree | - Strongly disagree | - Neither agree nor disagree | - Agree | - Strongly agree |
| Q7 | Working with people with lived experience is not part of my role as a researcher | | | | |
|  | - Strongly disagree | - Strongly disagree | - Neither agree nor disagree | - Agree | - Strongly agree |
| Q8 | Working with people with lived experience would be easy | | | | |
|  | - Strongly disagree | - Strongly disagree | - Neither agree nor disagree | - Agree | - Strongly agree |
| Q9 | Working with people with lived experience would not be of benefit | | | | |
|  | - Strongly disagree | - Strongly disagree | - Neither agree nor disagree | - Agree | - Strongly agree |
| Q10 | Working on a research project with people with lived experience would make me feel uncomfortable | | | | |
|  | - Strongly disagree | - Strongly disagree | - Neither agree nor disagree | - Agree | - Strongly agree |

**These questions are specifically about the training module you have just completed.**

| Q11 | Please rate your satisfaction with the content of the training module | | | | | | | | | |
| --- | --- | --- | --- | --- | --- | --- | --- | --- | --- | --- |
|  | ❑ 1 | ❑ 2 | ❑ 3 | ❑ 4 | ❑ 5 | ❑ 6 | ❑ 7 | ❑ 8 | ❑ 9 | ❑ 10 |
|  | Not at all satisfied | |  |  |  |  |  |  | Extremely satisfied | |
| Q12 | Please rate your satisfaction with the way the content was presented | | | | | | | | | |
|  | ❑ 1 | ❑ 2 | ❑ 3 | ❑ 4 | ❑ 5 | ❑ 6 | ❑ 7 | ❑ 8 | ❑ 9 | ❑ 10 |
|  | Not at all satisfied | |  |  |  |  |  |  | Extremely satisfied | |

| Q13 | Please rate how useful you found the training module for informing your research practice | | | | | | | | | |
| --- | --- | --- | --- | --- | --- | --- | --- | --- | --- | --- |
|  | ❑ 1 | ❑ 2 | ❑ 3 | ❑ 4 | ❑ 5 | ❑ 6 | ❑ 7 | ❑ 8 | ❑ 9 | ❑ 10 |
|  | Not at all useful | |  |  |  |  |  |  | Extremely useful | |
| Q14: | Would you recommend this training module to other researchers in the stroke field?  ❑ Yes ❑ No | | | | | | | | | |
| Q15 | Would you like further training on this topic?  ❑ Yes (explain what further training you would like):  ❑ No | | | | | | | | | |
| Q16 | Do you have any other comments about this training module? | | | | | | | | | |

| Q17 | Are you willing to participate in an interview about your experiences with the training module and involving people with lived experience in your research?  If so, please provide your contact details  Name:  Email address: |
| --- | --- |

**Supplementary File 2: Interview Topic Guides**

People with Lived Experience

1.	Please describe your experience of working with researchers on research projects

2.	What motivates you to work with researchers on research projects?

3.	What impact do you think that involvement of people with lived experience brings to research?

4.	Has your thinking or feeling changed since completing this training module?

5.	Are you more likely to seek to be involved in a research project having completed this training?

•	If so, how?

•	If not, why?

6.	What were the most useful pieces of the training package for you?

•	Why was it useful

7.	Is there anything that you would have liked to have seen included in this training that wasn’t?

•	What could be enhanced in the training package?

8.	Was there anything in this training package that will change the way you work with researchers?

•	If so, how?

•	How do you think that you could apply what you learned to an upcoming project?

9.	Are there any outstanding barriers to you being involved in a research project?

•	What do you think would address these outstanding barriers?

Prompt – support/mentoring/training for people with lived experience, support/mentoring/training for researchers etc.

10.	Anything else?

Academic Researchers

1.	Please describe your experience of working with people with lived experience.

•	What do you feel about including people with lived experience in your research?

•	How do you feel about including people with lived experience with post-stroke cognitive and communication difficulties in your research?

2.	What motivates you to include people with lived experience in your research? [motivation]

3.	What impact do you think that involving people with lived experience brings to research?

4.	Has your thinking or feeling changed since completing this training module?

5.	Are you more likely to involve a person with lived experience in your research project having completed this training?

•	If so, how?

•	If not, why?

6.	What were the most useful pieces of the training package for you?

•	Why was it useful

7.	Is there anything that you would have liked to have seen included in this training that wasn’t?

•	What could be enhanced in the training package?

8.	Do you think that you will implement any of the information and knowledge that you learned in this training package into your research project?

•	If so, how?

•	How do you think that you could apply what you learned to an upcoming project?

9.	Are there any outstanding barriers to involving a person with lived experience in your research?

•	What do you think would address these outstanding barriers?

•	Prompt – supervision / mentoring / more training etc.

10.	Anything else?
